# Long-term Graduate Outcomes of the First Integrated Bachelor of Medicine (MB)/Doctor of Philosophy (PhD) Programme in Europe

**DOI:** 10.1101/2025.07.24.25331924

**Authors:** Ardon M Pillay, Hannah Dennis, Robert K. Semple, Stefan J Marciniak, D Keith Peters, Timothy M Cox

**Affiliations:** School of Clinical Medicine, University of Cambridge, Cambridge UK; Department of Pathology, University of Cambridge, Cambridge, UK; Centre for Cardiovascular Science, University of Edinburgh, Edinburgh, UK; MRC Human Genetics Unit, University of Edinburgh, Edinburgh, UK

## Abstract

**Background:** The Cambridge Bachelor of Medicine (MB)/Doctor of Philosophy (PhD) programme was established in 1989 as Europe’s first integrated clinical -doctoral research training programme. Here we evaluate long-term outcomes of the programme 36 years after its introduction.

**Methods:** To track mature career choices, research productivity, group leader and senior authorship status, we analysed the career outcomes of graduates enrolling on the programme between 1989 and 2014. This was accomplished by scrutinising institutional pages, LinkedIn, ResearchGate, Google Scholar, and PubMed profiles of MB/PhD programme alumni.

**Results:** We obtained data for 166 of 183 programme alumni (91%) who enrolled between 1989 and 2014, with a median follow up of 16 years. Among graduates, 139 (84%) remain in clinical practice and 27 (16%) transitioned to non-clinical careers. Clinical graduates entered medical (81%) and surgical (19%) specialties, with neurology and medical oncology most frequently chosen among the medical specialties. Neurosurgery and ophthalmology were the most favoured surgical specialties chosen. Research engagement remains high: 135 graduates (81%) have published research within the last five years and 28 (17%) hold group leadership positions. Of the the graduates no longer in clinical practice, most have taken up positions in industry (59%) and almost one fifth remain in academia.

**Conclusions:** The Cambridge MB/PhD programme has produced versatile graduates who contribute to the advancement of medicine within its complex professional ecosystem. Active research engagement (81%) and securement of group leadership positions among alumni (17%), attest to the ability of our graduates to sustain academic career choices within the context of contemporary clinical practice. That most graduates also continue to conduct research shows that the original goals of the programme have been met.

## Introduction

The medical profession has a responsibility not only to treat illness and prevent disease but to introduce and advance knowledge on which these responsbilities depend. Academic medicine, in whatever guise, is the natural career path chosen by clinical students who are able and sufficiently motivated to seek the scientific training needed to conduct innovative medical research.

During a period of disruptive changes in medical education across the globe, much has been written about the need to develop academic medicine within universities^1–4^. Mindful of concerns about the future of biomedical research and the ability of medical undergraduates and young physicians to engage with scientific advances, the University of Cambridge introduced an integrated programme that provides training in both research and clinical medicine^5,6^. This course is offered as an alternative to the traditional clinical training course, which comprises a three-year undergraduate bachelor’s degree in medical sciences (BA Hons) followed by three years of clinical training to obtain the qualifying Bachelor’s degrees in medicine (MB) and surgery (BChir). Those undertaking the MB/PhD (Doctor of Philosophy) programme complete one year of clinical training after completing their bachelor’s degree before undertaking an intercalated PhD (Figure 1). This provides an early career stage opportunity for training in scientific research. Postgraduate fees are paid by the MB/PhD programme at the UK home rate; overseas student fees are found by the student (often through Cambridge Overseas Trusts). During the PhD phase, students are offered a minimum stipend of £22,280 rising to 23,320 for three research years (2025-27 rates) from institutional funds. Modest consumables (£2500 pa) are also provided to the hosting laboratory group. To maintain core clinical skills during the intercalated PhD, students are required to attend regular clinical teaching sessions. On completing their PhD, they return to full-time clinical training for two years.

**Figure 1:**
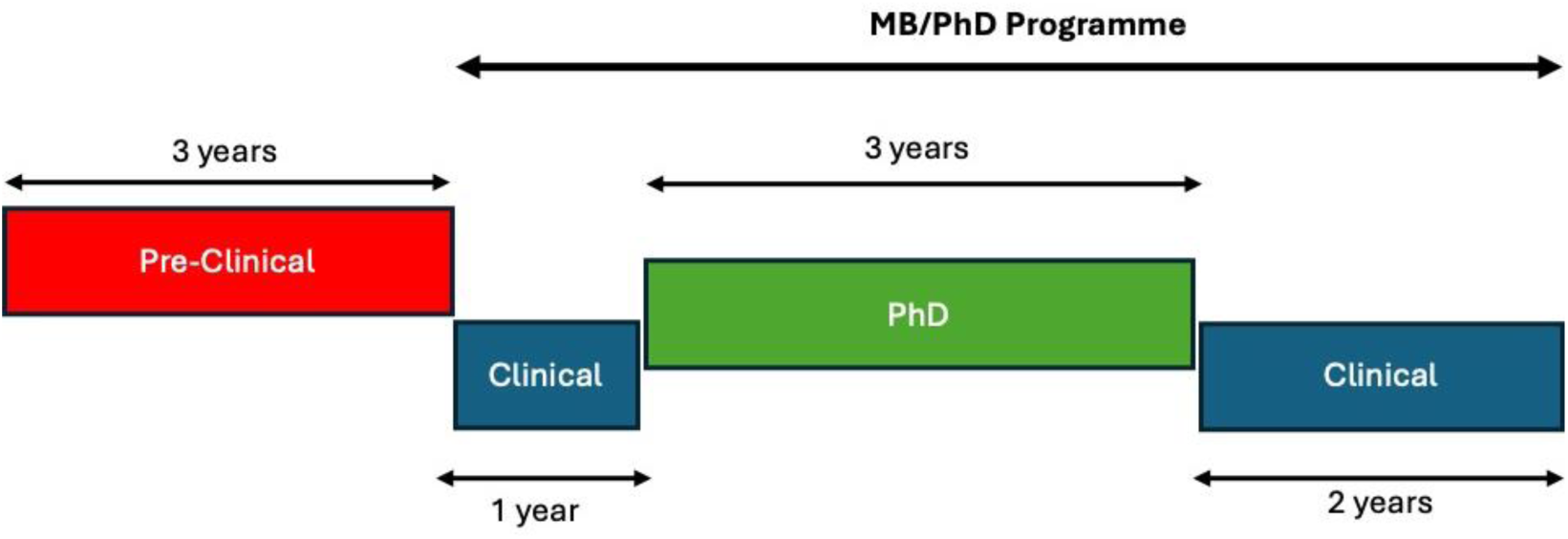
The timeline of the Cambridge MB/PhD programme. MB/PhD students, after the completion of the pre-clinical course (3 years) and first year of clinical medicine, undertake an intercalated PhD. When the PhD has been completed, they return to their final two years of clinical training.

After the Cambridge MB/PhD programme was established, similar programmes were set up at other British medical schools, including University College London in 1994^7^ and Imperial College London^8^, followed rapidly by others^9^. Thirty six years after its inception, we review the Cambridge MB/PhD programme using new data on graduate outcomes. The results and implications of this exploratory work are set out here.

## Methods

To examine career paths at least five years after completion of the MB BChir (and PhD), we focused on alumni who had completed their courses of study and had been awarded the PhD and MB BChir degrees by 2020. We selected this timeframe as this set of alumni will have had at least five years after clinical qualification to undertake specialty training. Using institute or company biographical pages, LinkedIn, PubMed, ResearchGate, and Google Scholar, we identified graduate career paths and clinical specialties. We isolated data for 166 (91%) of the 183 alumni who completed the full programme by the end of 2020. The follow up time after completion of the programme ranged from five to thirty years (median 16 years). The information enabled us to address the nine questions set out below.

1. Are they still practising medicine?
2. Are they in a medical or surgical specialty?
3. In what specialty within medicine or surgery are they employed?
4. Have they published academic articles during the last five years?
5. Has any academic work published in the last five years utilised human materials?
6. Are they group leaders?
7. Are they senior authors on academic publications that have appeared in the last five years?
8. If they are not practising medicine, in which fields are they employed?
9. In which country do they currently work ?

Records of programme members were accessed minimally by the authors to ensure that data and figures presented are confidential and non-identifiable.

## Results

### Conspectus of the MB/PhD programme

Since its inception in 1989, when a single student enrolled, to the 35^th^ anniversary, 258 students joined the Cambridge MB/PhD programme, of whom 98 (38%) were women. The proportion of women who enroll on the programme has varied considerably (Supplementary Figure 1); at the time of data collection, of the 46 MB/PhD students enrolled on the programme, 21 are women (46%) (Table 1).

**Table 1.** Current Status of the Cambridge MB/PhD Programme.

| Type of Student | Number of Students | Number of Women | Number of Men |
| --- | --- | --- | --- |
| Alumni | 212 | 77 (36%) | 135 (64%) |
| Current Students | 46 | 21 (46%) | 25 (54%) |
| Total | 258 | 98 (38%) | 160 (62%) |

Two hundred and twelve MB/PhD students have formally left the programme. Of these 212 alumni, 8 (4%) obtained their MB/BChir degree but did not complete their PhD, 6 (3%) completed their PhD but did not receive their MB/BChir degree; 2 (<1%) did not obtain either degree (Table 2). The mean duration until the award of the PhD degree was < 4 years (n=204).

**Table 2.** Cambridge MB/PhD Programme Alumni.

| Classification | Number | Percentage of Alumni (%) |
| --- | --- | --- |
| Alumni | 212 | 100 |
| Did not complete PhD | 8 | 4 |
| Did not complete MB/BChir | 6 | 3 |
| Did not complete PhD or MB/BChir | 2 | <1 |

### An overview of MB/PhD graduate outcomes

Of the 166 identified alumni who completed the full programme by the end of 2020, 139 (84%) graduates continue to work in medical or surgical specialties (Table 3). Among these, 119 alumni (86%) have published in the scientific literature in the past five years, 27 (19%) hold group leader positions, 18 (13%) are senior authors on recent publications (Figure 2). Twenty seven graduates (16%) no longer practise medicine. Overall, 135 graduates (81%) have published in the last five years and 28 (17%) are group leaders. Among the former, two thirds have conducted research on persons or human tissues at least once in the past five years.

**Figure 2.**
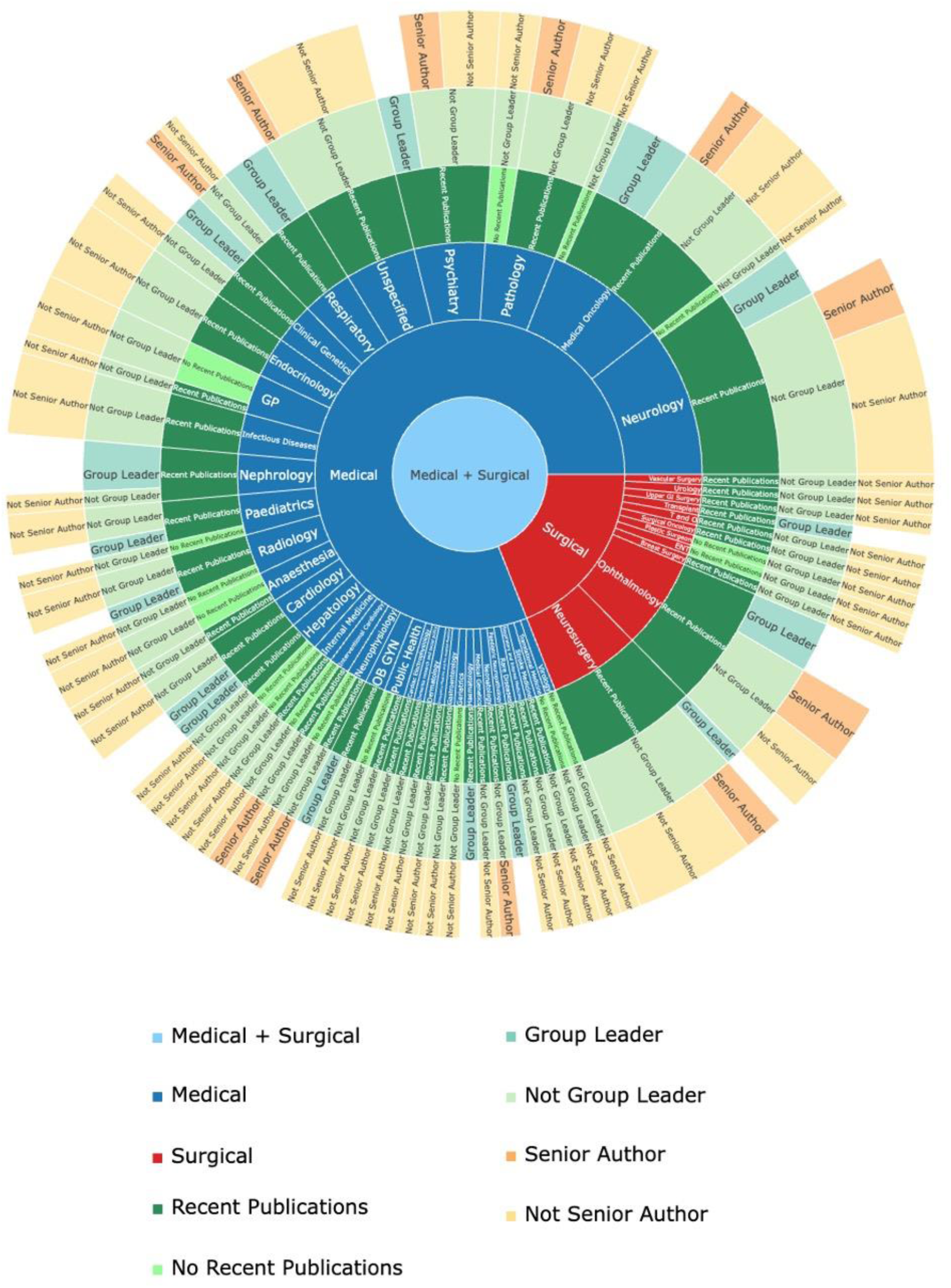
A sunburst chart showing career outcomes of MB/PhD graduates (1989-2014) who are in medical practice. The chart indicates if they are practising in a medical or surgical specialty; in which specialty they practise; if they have published academic research in the last five years; if they are group leaders and if they have been senior authors on any academic research they have published in the last five years. OB GYN = Obstetrics and Gynaecology, ENT = Ear, Nose, and Throat Surgery, T&O = Trauma and Orthopaedics, Unspecified = specialty undetermined.

**Table 3:** Overview of MB-PhD Graduate Outcomes for 166 of 183 Alumni (91%)

| Classification | Number of Graduates |
| --- | --- |
| Alumni identified | 166 |
| Medical/surgical specialists | 139 (84%) |
| Not in clinical practice | 27 (16%) |
| Published (past five years) | 135 (81%) |
| Group leaders | 28 (17%) |
| Senior authors | 18 (13%) |

Of those who continue to practise clinical medicine, 112 graduates are employed in medical specialties (81%) and 27 in surgical specialties (19%). In the past five years, 94 alumni in medical specialties (84%) have published whereas 25 of those within surgical specialties (94%) have research publications.

Among graduates practising in medical specialties, 13 (12%) have been senior authors in the last five years. The equivalent number for those working in surgical specialties is 5 (19%).

### MB/PhD graduates in medical specialties

Graduates of the MB/PhD programme have taken up an extensive range of medical specialties, including Psychiatry, Dermatology, and Emergency Medicine (Figure 3 and Table 4).

**Figure 3.**
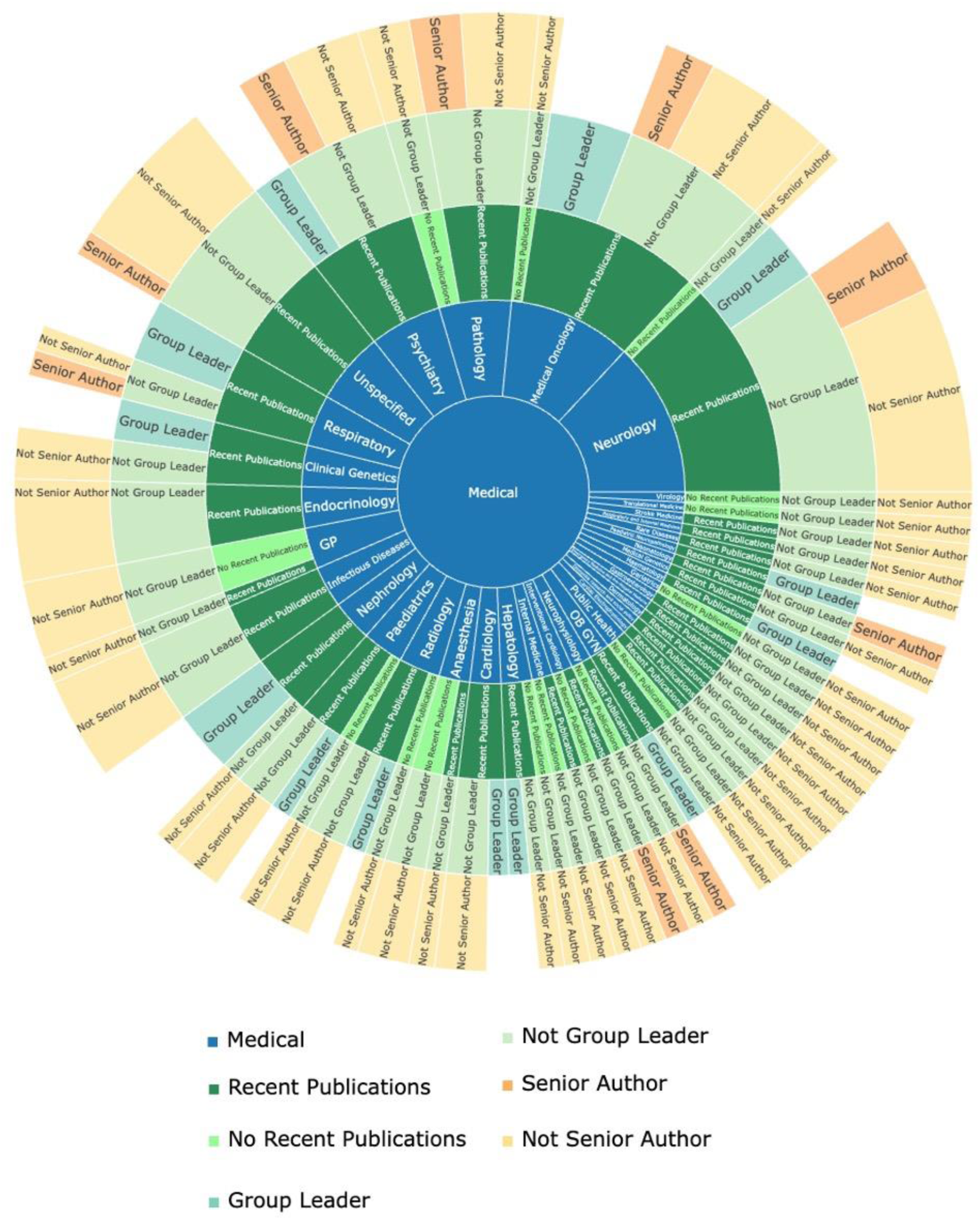
A sunburst chart with career outcomes of MB/PhD graduates (1989-2014) practising clinical medicine. The chart shows the specialty in which they practise; if they have published academic research in the last five years, if they are group leaders, and if they have been senior authors on any academic research they have published in the last five years. OB GYN = Obstetrics and Gynaecology, Unspecified = specialty undetermined.

**Table 4.**
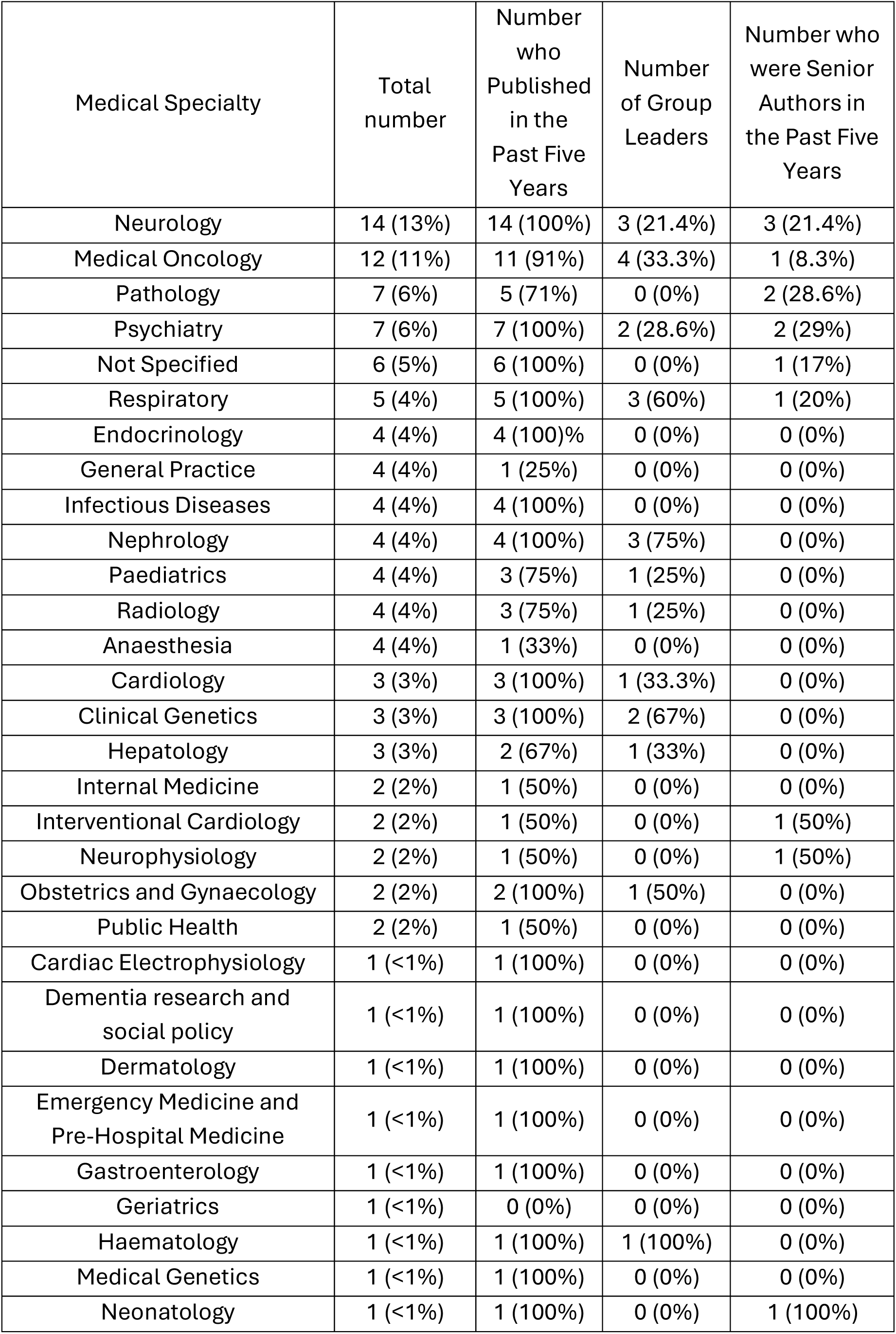
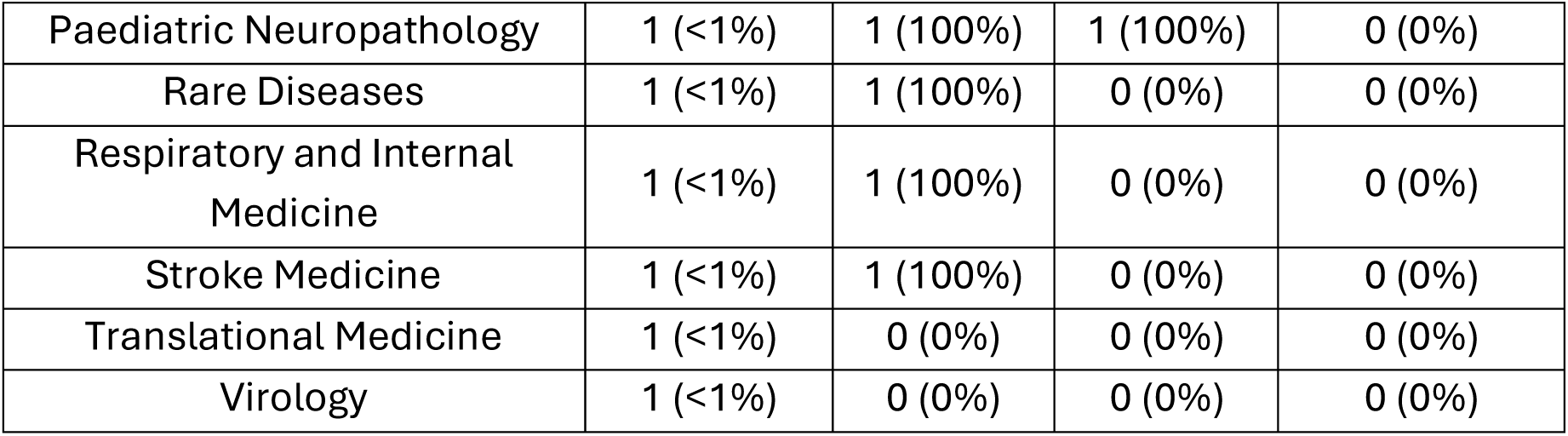
Cambridge MB/PhD Graduates in Medical Specialties.

| Medical Specialty | Total number | Number who Published in the Past Five Years | Number of Group Leaders | Number who were Senior Authors in the Past Five Years |
| --- | --- | --- | --- | --- |
| Neurology | 14 (13%) | 14 (100%) | 3 (21.4%) | 3 (21.4%) |
| Medical Oncology | 12 (11%) | 11 (91%) | 4 (33.3%) | 1 (8.3%) |
| Pathology | 7 (6%) | 5 (71%) | 0 (0%) | 2 (28.6%) |
| Psychiatry | 7 (6%) | 7 (100%) | 2 (28.6%) | 2 (29%) |
| Not Specified | 6 (5%) | 6 (100%) | 0 (0%) | 1 (17%) |
| Respiratory | 5 (4%) | 5 (100%) | 3 (60%) | 1 (20%) |
| Endocrinology | 4 (4%) | 4 (100%) | 0 (0%) | 0 (0%) |
| General Practice | 4 (4%) | 1 (25%) | 0 (0%) | 0 (0%) |
| Infectious Diseases | 4 (4%) | 4 (100%) | 0 (0%) | 0 (0%) |
| Nephrology | 4 (4%) | 4 (100%) | 3 (75%) | 0 (0%) |
| Paediatrics | 4 (4%) | 3 (75%) | 1 (25%) | 0 (0%) |
| Radiology | 4 (4%) | 3 (75%) | 1 (25%) | 0 (0%) |
| Anaesthesia | 4 (4%) | 1 (33%) | 0 (0%) | 0 (0%) |
| Cardiology | 3 (3%) | 3 (100%) | 1 (33.3%) | 0 (0%) |
| Clinical Genetics | 3 (3%) | 3 (100%) | 2 (67%) | 0 (0%) |
| Hepatology | 3 (3%) | 2 (67%) | 1 (33%) | 0 (0%) |
| Internal Medicine | 2 (2%) | 1 (50%) | 0 (0%) | 0 (0%) |
| Interventional Cardiology | 2 (2%) | 1 (50%) | 0 (0%) | 1 (50%) |
| Neurophysiology | 2 (2%) | 1 (50%) | 0 (0%) | 1 (50%) |
| Obstetrics and Gynaecology | 2 (2%) | 2 (100%) | 1 (50%) | 0 (0%) |
| Public Health | 2 (2%) | 1 (50%) | 0 (0%) | 0 (0%) |
| Cardiac Electrophysiology | 1 (<1%) | 1 (100%) | 0 (0%) | 0 (0%) |
| Dementia research and social policy | 1 (<1%) | 1 (100%) | 0 (0%) | 0 (0%) |
| Dermatology | 1 (<1%) | 1 (100%) | 0 (0%) | 0 (0%) |
| Emergency Medicine and Pre-Hospital Medicine | 1 (<1%) | 1 (100%) | 0 (0%) | 0 (0%) |
| Gastroenterology | 1 (<1%) | 1 (100%) | 0 (0%) | 0 (0%) |
| Geriatrics | 1 (<1%) | 0 (0%) | 0 (0%) | 0 (0%) |
| Haematology | 1 (<1%) | 1 (100%) | 1 (100%) | 0 (0%) |
| Medical Genetics | 1 (<1%) | 1 (100%) | 0 (0%) | 0 (0%) |
| Neonatology | 1 (<1%) | 1 (100%) | 0 (0%) | 1 (100%) |
| Paediatric Neuropathology | 1 (<1%) | 1 (100%) | 1 (100%) | 0 (0%) |
| Rare Diseases | 1 (<1%) | 1 (100%) | 0 (0%) | 0 (0%) |
| Respiratory and Internal Medicine | 1 (<1%) | 1 (100%) | 0 (0%) | 0 (0%) |
| Stroke Medicine | 1 (<1%) | 1 (100%) | 0 (0%) | 0 (0%) |
| Translational Medicine | 1 (<1%) | 0 (0%) | 0 (0%) | 0 (0%) |
| Virology | 1 (<1%) | 0 (0%) | 0 (0%) | 0 (0%) |

The most represented specialties among these graduates are Neurology (13%), Medical Oncology (11%), and Pathology (6%). Four graduates have entered General Practice. As mentioned, 84% of graduates working in medical specialties continue to publish their research. Within certain medical specialties (namely Neurology, Psychiatry, Respiratory,

Endocrinology, Infectious Diseases, Nephrology, Cardiology, Clinical Genetics), every identified graduate has produced research publications in the past five years.

### MB/PhD graduates in surgical specialties

The 27 MB/PhD graduates who chose surgical careers also joined a range of specialties (Figure 4 and Table 5). The modal specialties represented among graduates who became surgeons were Neurosurgery (33%) and Ophthalmology (33%).

**Figure 4.**
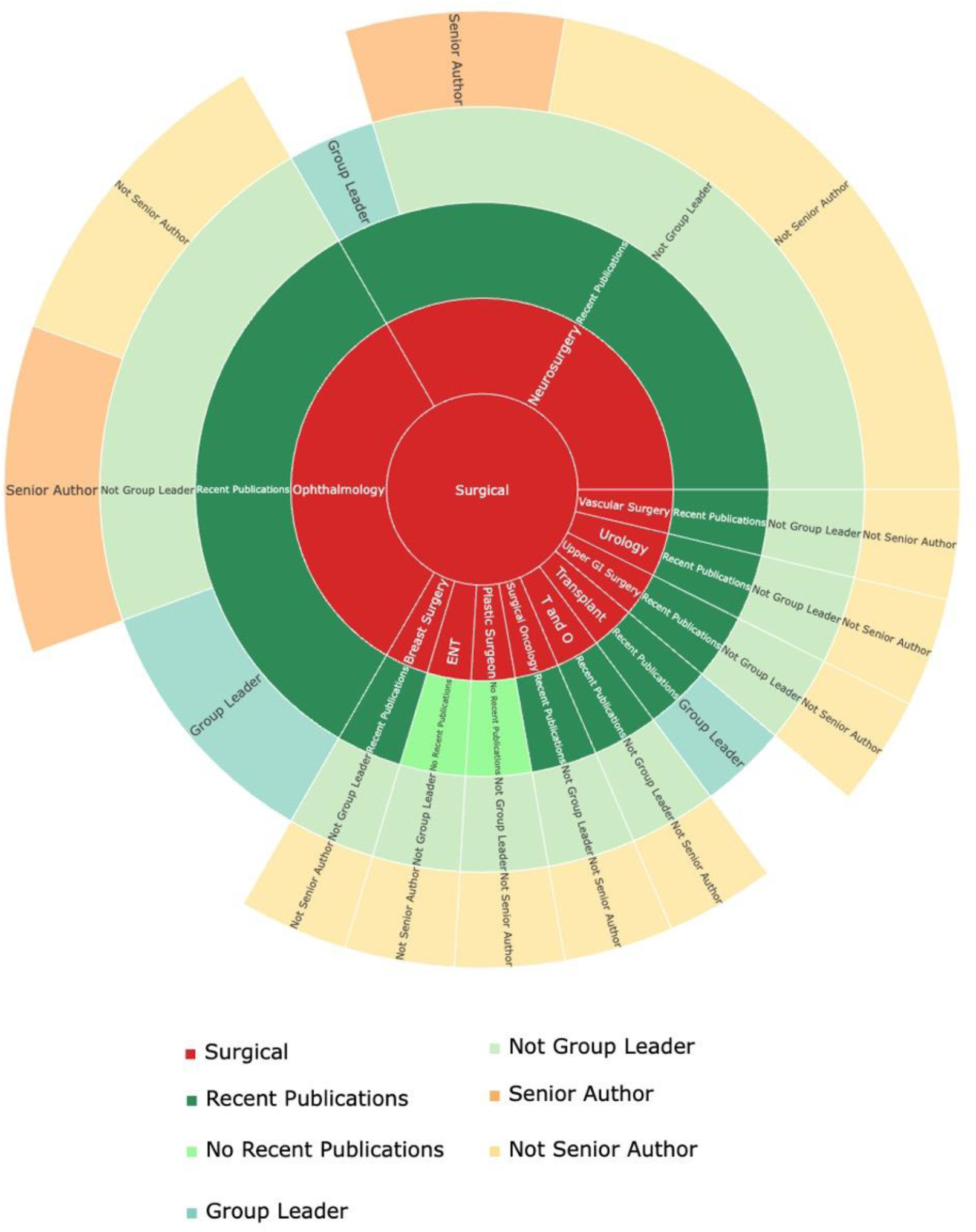
A sunburst chart with career outcomes of MB/PhD graduates (1989-2014) practising in surgical specialties. The chart shows the specialty in which they practise; if they have published academic research in the last five years; if they are group leaders and if they have been senior authors on any academic research they have published in the last five years. ENT = Ear, Nose, and Throat Surgery, T&O = Trauma and Orthopaedics.

**Table 5.** Cambridge MB/PhD Graduates in Surgical Specialties.

| Surgical Specialty | Total Number | Number who Published in the Past Five Years | Number of Group Leaders | Number who were Senior Authors in the Past Five Years |
| --- | --- | --- | --- | --- |
| Neurosurgery | 9 (33%) | 9 (100%) | 0 (0%) | 2 (22%) |
| Ophthalmology | 9 (33%) | 9 (100%) | 2 (22%) | 3 (33%) |
| Breast Surgery | 1 (4%) | 1 (100%) | 0 (0%) | 0 (0%) |
| Ear, Nose, and Throat Surgery | 1 (4%) | 0 (0%) | 0 (0%) | 0 (0%) |
| Plastic Surgeon | 1 (4%) | 0 (0%) | 0 (0%) | 0 (0%) |
| Surgical Oncology | 1 (4%) | 1 (100%) | 0 (0%) | 0 (0%) |
| Trauma and Orthopaedics | 1 (4%) | 1 (100%) | 0 (0%) | 0 (0%) |
| Transplant | 1 (4%) | 1 (100%) | 1 (100%) | 0 (0%) |
| Upper GI Surgery | 1 (4%) | 1 (100%) | 0 (0%) | 0 (0%) |
| Urology | 1 (4%) | 1 (100%) | 0 (0%) | 0 (0%) |
| Vascular Surgery | 1 (4%) | 1 (100%) | 0 (0%) | 0 (0%) |

Readers are invited further to explore the data relating to each medical and surgical specialty by means of the interactive version of figure 2 (Supplementary Data File 1).

### Graduates No Longer in Clinical Practice

Twenty seven MB/PhD graduates no longer practise medicine (19%) and have either continued in academia, pursued careers in industry or undertaken miscellaneous roles (Figure 5). Of the 27 individuals, 16 have published scientific articles in the last five years (59%).

**Figure 5:**
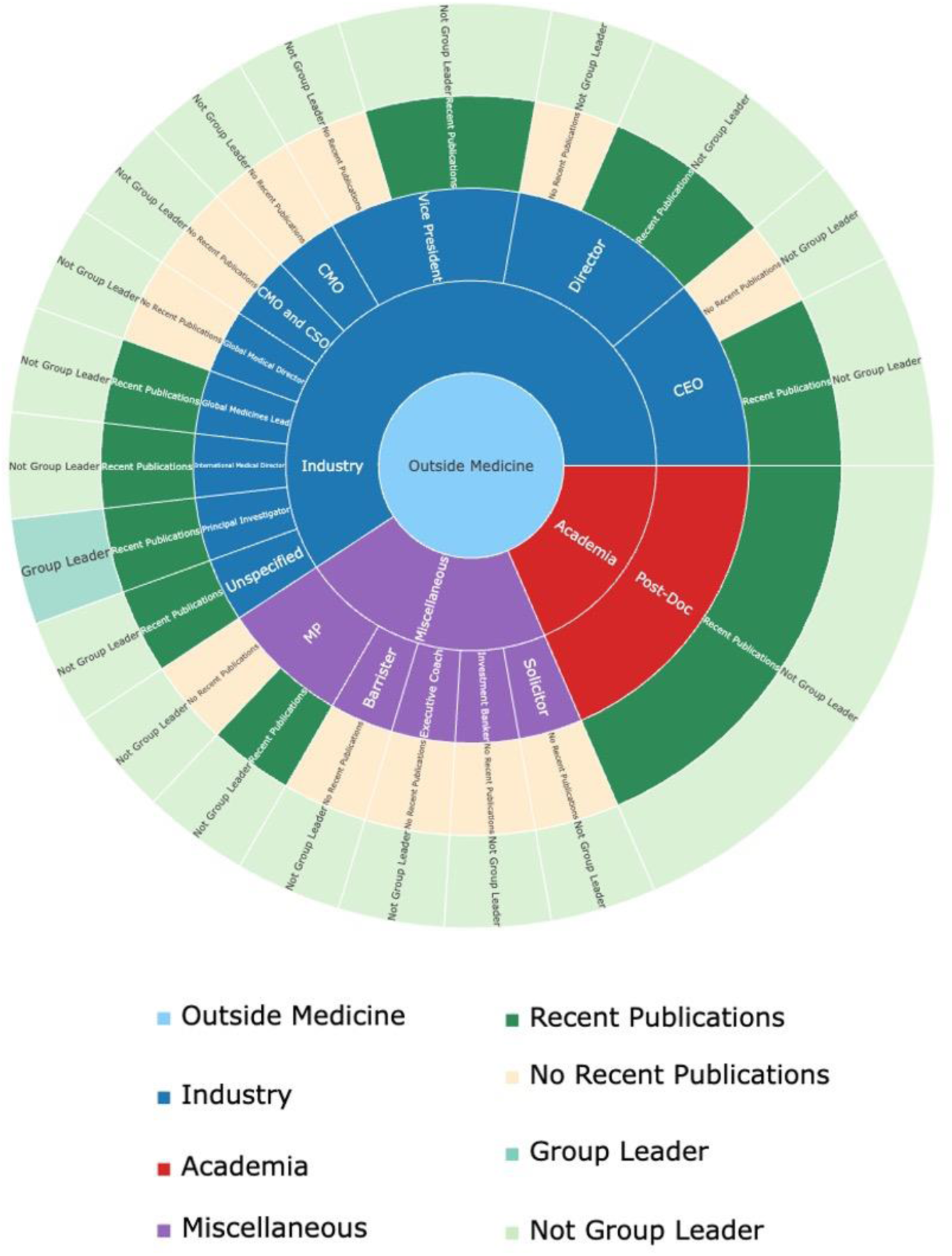
A sunburst chart with graduate outcomes of MB/PhD graduates (1989-2014) who do not currently practice medicine. The chart indicates the nature of their current careers, their current titles, if they have published academic research in the last five years, and if they are group leaders. MP = member of parliament, CEO = chief executive officer, CMO = chief medical officer, CSO = chief scientific officer.

Industry has been the most favoured alternative career (16 graduates, 59%) among graduates who are no longer in clinical practice. Those employed in industry have diverse roles, including positions in senior management and as principal investigators. Five of the 27 non-practising alumni are full-time academics (19%). Six graduates have moved into careers with limited apparent connections to clinical medicine or science. Two of these graduates are members of parliaments; the remainder are a solicitor, a barrister, an investment banker and an executive coach.

The reader is invited to examine these data further by using the interactive version of figure 5 (Supplementary Data File 2).

#### Geographical Distribution of Graduates

We were able to determine the countries of residence for 160 of the 166 identified alumni who completed the full programme by the end of 2020 (96%): 129 (81%) alumni remain in the United Kingdom (UK), 15 (9%) are based in the United States of America (USA), 13 (8%) reside in Singapore, and 3 (<1%) others reside in other countries.

## Discussion

Almost immediately after its introduction, the Cambridge MB/PhD programme attracted considerable controversy: some respected local clinical teachers vehemently opposed this new model^5,10^. Thirty six years after its inception, graduates of the programme contribute to numerous fields of endeavour across medicine. Many alumni also occupy creative roles in medical education and drug development. Notably, their employment is not geographically restricted to the UK – programme alumni also work in the USA, Singapore, and other countries.

From the outset, we recognised that radical changes in the practice of medicine and especially the organization of clinical higher medical training were rapidly being introduced, alongsidechanges in the universities and organisation of the National Health Service^5^. All these factors impinge on those who aspire to pursue a research-based career^5^. Opportunities to secure early scientific training, before the constraints of professional employment and personal responsibilities grow, may mitigate the reported high attrition rate in the conventional academic medicine career pathway^11^. In this regard, we note that women have continued to join the programme throughout its history (supplementary Figure 1). Across the thirty six year history of the MB/PhD programme, 38% of those enrolled have been women and, among the current body of programme members, nearly half are women (Table 1).

The structure of the programme (Figure 1) enables the early acquisition of research skills and experience by postgraduates who are already fully engaged as students of clinical medicine. Early conduct of research was found to be a strong determinant of progress in clinical scientific careers that lead to tenure-track positions as principal investigators^9^. This is especially relevant, given that 28 of 166 identified alumni are already group leaders. Furthermore >80% of these alumni are research active, which confirms their vested interest in, and commitment to continuing investigative work in the long term.

MB/PhD alumni who are practising physicians have joined a large assortment of specialties, from Neurology to General Practice (Figure 2). This range of occupations illustrates how undertaking an immersive period of research early in one’s career does not materially restrict professional opportunities ahead.

Academic surgery provides a vivid example of how integrated MB/PhD programmes can favour future career development. The fundamental nature of surgical training and practice is an extreme example of the tension that stems from the practical conflict between the need for protected time in research alongside training demands for hands-on experience in surgical practice. Surgical trainees who take time out to undertake a PhD may have reduced clinical hours, thus further prolonging the time needed to meet mandatory competency requirements. The MB/PhD programme allows aspiring surgeons to develop key scientific skills within at least 3 years of protected time before barriers to achieving sufficient clinical and research competencies present themselves later in training^12,13^, with the opportunity to achieve greater scientific prowess during surgical training and practice than their full-time surgical peers. This observation is supported by the finding that > 90% of MB/PhD graduates working in surgical specialties are active in research and have authored publications in the last five years (Figure 4). However, publications from some surgeons include case reports, a publication form which is often less dependent on research training in science but equally may inspire new projects.

The general conclusion is that there is an enduring and high level of engagement in research among the MB/PhD alumni. This, combined with the impressive proportion of group leaders emerging from the programme (17%), signals the effective development of committed physician-scientists, as was envisaged. We thus contend that there is a clear role for similar integrated research training programmes within contemporary medical schools. Such programmes will contribute to the expansion and development of a cadre of investigators at an early stage of their career who are anchored within real-world clinical practice and thus invaluable for the advancement of medicine.

Notably, not all MB/PhD alumni pursue traditional clinical academic careers. Among the 27 graduates who moved away from clinical medicine, most continue to conduct research (Figure 5). Many of these individuals contribute meaningfully to medical advancement through non-traditional but nonetheless familiar pathways. Those working in biotechnology and pharmaceutical companies (nearly two-thirds of those no longer working in clinical medicine) seek to introduce innovative technologies and therapies with direct clinical applications^14,15^. Such work requires scientific rigour and a practical understanding of clinical realities – abilities that are fostered by truly integrated MB/PhD programmes.

The scientific skills developed through our training programme thus extend beyond traditional academic medicine. The programme places emphasis on critical thinking and problem-solving - abilities that promote the advancement of medicine and beyond. The versatility of these skills is exemplified in those MB/PhD graduates who have moved successfully into finance, government and the law - domains of human endeavour to which their combination of medical knowledge and scientific expertise brings a unique perspective.

The breadth of career paths pursued by MB/PhD graduates can be viewed as a mark of success of this programme in developing adaptable professionals who contribute to society in many ways, whether by advancing medical science directly or applying their expertise more broadly.

The MB/PhD programme contributes to the diversity of opportunities for clinician-scientists and is now a sustained and successful training option. Given the cost disparity between support for MB/PhD programme students and clinical training fellowships and related research schemes for qualified clinicians, the programme appears to be an efficient and economical means to generate academic clinicians. A further potential advantage of early exposure to contemporary scientific research is that younger people tend to have an aptitude for fundamental methodologies - to which, on account of the distractions of speciality training, those supported by clinical fellowships are likely to be more adverse.

Our analysis lacks a direct control group to ascertain the extent to which the graduate outcomes can be attributed to the programme itself, rather than the characteristics of those who applied for acceptance. However, this study is intended solely to report and comment on the mature outcomes, without dogmatic assertions that the programme alone was responsible for the careers secured by alumni. Members of the programme are selected from a self-chosen group of applicants. Thus the appropriate control group would include those accepted for admission but who for one reason or another did not join the programme. There are very few such individuals. A less appropriate but general control group would be clinical undergraduates who did not apply for the programme. Clearly many such persons would be suitably qualified.

Taken together, we argue that the Cambridge MB/PhD programme has produced alumni who are well placed to contribute to the development of academic medicine at several levels. Among the alumni are politicians who allocate resources for healthcare; group leaders, who enhance links between core and clinical biomedical science; scientist, who investigate fundamental biological questions; researchers in industry, who develop new therapeutics and clinician-scientists who break new ground on hitherto unknown aspects of human disease .

These graduates personify the founding goal of the Cambridge MB/PhD programme – the development of individuals with an investigative nature and advanced scientific training, ultimately enabling them to translate original research into tangible benefits for society.

## Data Availability

All data produced in the present study are confidential.

## Acknowledgements

We thank Professor Paul Wilkinson (Clinical Dean) and Professor Patrick Maxwell CBE, Regius Professor of Physic and over many years, clinical school staff for their continuing support of the programme.

## Competing Interests

### No conflicts

A.M.P is a current student enrolled on the Cambridge MB/PhD programme. S.J.M currently directs the Cambridge MB/PhD programme and is one of its alumni. R.S and T.M.C have previously directed the programme. All other authors are, or have been directly involved with the inception, establishment, direction, management, administration and/or fund-raising related to the MB/PhD programme at the University of Cambridge.

## Supplementary Material

**Supplementary Figure 1:**
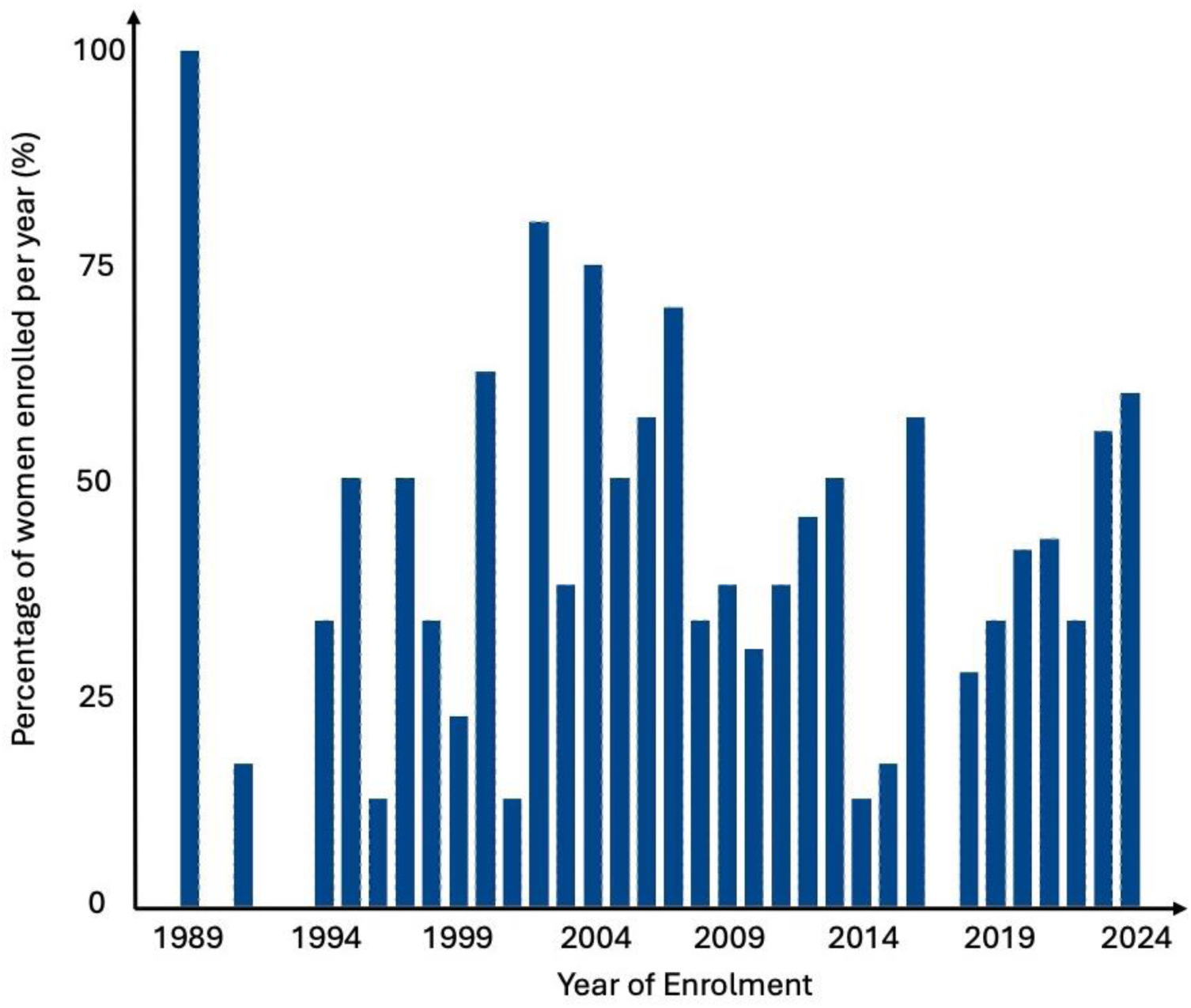
A bar graph showing the percentage of women enrolled on the MB/PhD women in every intake year (1989-2024).

**Supplementary File 1: An interactive version of figure 1 in .html format. Please download and open in your web browser to use the interactive features.**

**Supplementary File 2: An interactive version of figure 4 in .html format. Please download and open in your web browser to use the interactive features.**

